# Systematic review of the evidence of cardiac dysfunction and intravenous fluid use among children with severe acute malnutrition and severe dehydration

**DOI:** 10.1101/2025.08.20.25333892

**Authors:** Rupa Narra, Kathryn Alberti, Iza Ciglenecki, Ben GS Allen, Kirrily de Polnay, Kirkby D Tickell

## Abstract

**Background:** A sizeable proportion of cholera attributed mortality occurs among children with severe acute malnutrition (SAM). International guidelines severely limit the use of intravenous fluids among children with SAM and cholera due to their perceived vulnerability to fluid overload. We reviewed the cardiac function evidence and studies of intravenous fluid use among children with SAM.

**Methods:** Two systematic reviews of English language papers published after the last Global Task Force for Cholera Control recommendations on the treatment of children with cholera and SAM (2017) were conducted. The first included studies of cardiac function among children presenting to hospital with SAM. The second included studies of intravenous fluid use among children with dehydrating diarrhea and SAM.

**Results:** Among eight echocardiographic studies of cardiac function in children with SAM at presentation to hospital, all found left ventricular mass to be decreased but none found systolic function to be impaired. Seven of the eight studies assessing diastolic function found some degree of mild-to-moderate impairment. Two echocardiographic studies monitored responsiveness to intravenous fluid resuscitation and found no evidence of fluid overload. Finally, three studies gave intravenous fluids to children with SAM and either shock or dehydration. Collectively, these studies included 117 children with SAM given intravenous fluid. No cases of fluid overload were observed.

**Conclusion:** No evidence to support decreased systolic function among children with SAM and no documented cases of fluid overload during intravenous rehydration of these children were identified. These data call into question guidelines that severely restrict intravenous fluids among children with SAM and highly dehydrating conditions such cholera.

## Introduction

Cholera is a diarrhoeal disease caused by the bacteria *Vibrio choler*ae and is estimated to affect up to four million people each year.[1] Since 2021, there has been a surge in cholera globally with several countries reporting the disease for the first time in decades. In 2023, the reported cholera deaths increased by over 70% compared to 2022, and 38% of cases were among children under five years of age.[2] Severe cholera is characterized by rapid fluid losses, adults can reach 1000 ml/hour, and if left untreated can have case fatality ratios of up to 50%.[3] Primary treatment for these patients is rapid intravenous rehydration, which should reduce case fatality rates to below 1%.[3]

WHO defines severe acute malnutrition (SAM) as a weight-for-height less than three standard deviations below the WHO child growth standards median (severe wasting); and/or a mid-upper arm circumference less than 115mm (for children 6-59 months); or the presence of nutritional oedema.[4] Children with SAM are at higher risk of death from any cause of diarrhea when compared to non-malnourished children,[5–8] but the management of children with SAM and comorbid cholera remains particularly challenging. Children with cholera often present with severe dehydration and/or hypovolemic shock,[9] and early administration of appropriate rehydration therapy is critical. However, WHO guidelines discourage intravenous rehydration among children with SAM because expert opinion suggested that these children had depressed cardiac function leaving them vulnerable to fluid overload.[10] In line with this expert opinion and the WHO guideline, the 2018 Global Task Force for Cholera Control (GTFCC, S1 Appendix 1) guidance reserves intravenous fluids for children with SAM and cholera and severe dehydration and altered mental status (lethargy or loss of consciousness) and those with ‘some dehydration’ to be treated with oral fluids only.[11] Since this guidance was reviewed in 2017, multiple publications have challenged the assumption that children with SAM systematically have underlying cardiac dysfunction and that they respond poorly to intravenous rehydration with a high risk of fluid overload leading to clinical deterioration and potentially death.

Recently published echocardiographic studies have suggested that children with SAM have normal systolic cardiac function for their body mass, indicating that they may tolerate, and benefit from, intravenous fluids when given for the appropriate clinical indication.[12–14] Several agencies have deviated from WHO and GTFCC guidance by managing these children with intravenous fluids if they have any form of dehydration. This includes Médecins Sans Frontières clinical guidelines, and clinical trials conducted by the international centre for diarrheal disease research Bangladesh (Icddr,b).[15,16] This review aims to summarize and critically appraise literature on the safety and efficacy of intravenous rehydration in children with SAM and cholera published since the *2018 GTFCC* recommendations. To better understand concerns of cardiac dysfunction and fluid overload among children with SAM we also reviewed echocardiographic studies among this population.

## Methods

There is limited literature on children with SAM and cholera, and therefore this review was expanded to children with SAM and any diarrheal disease complicated by some (sometimes referred to as moderate) or severe dehydration with or without hypovolemic shock. Two searches were conducted, the first identified echocardiography studies among children with SAM, and the second identified clinical studies of intravenous rehydration among children with SAM and dehydration with or without shock. Both searches were limited to studies published after the last GTFCC review period in 2017. SAM was defined as per WHO recommendations, weight-for-height (WHZ) or weight-for-length z-score (WLZ) z-score < -3, or mid-upper arm circumference (MUAC) < 115 mm (for children 6-59 months) and/or nutritional oedema.

Relevant literature published between January 1, 2017 - July 1, 2024 were identified and reviewed. Literature from clinical trials, meta-analyses, randomized control trials, observational studies, systematic reviews, grey literature, and guidance documents were eligible for inclusion. Data were collected from PubMed, MEDLINE, and Cochrane databases, non-governmental organizations and WHO websites. Search terms are listed in Table 1. Only literature published in English on human patients was included. Title abstract review was conducted by one author (RN), and eligibility confirmed by a second (KDT).

**Table 1.** Search terms, inclusion and exclusion criteria

|  | All searches | Search 1: Cardiac Function | Search 2: Intravenous rehydration |
| --- | --- | --- | --- |
| Topic | Both topics | Cardiac function in children with SAM | Children with SAM and diarrheal disease and severe dehydration with or without shock that received IV rehydration |
| Search terms | (‘Children’ OR ‘paediatric’) AND (‘malnutrition’ OR ‘marasmus’ OR ‘kwashiorkor’) | AND (‘cardiac’ OR ‘heart’ OR ‘myocardial’) AND (‘function’ OR ‘dysfunction’) | AND (‘cholera’ OR ‘acute watery diarrhoea’ OR ‘diarrhoea’ OR ‘diarrheal disease’) AND (‘dehydration’ OR ‘shock’ OR ‘rehydration’ OR ‘fluid therapy’ OR ‘intravenous hydration’) |
| Inclusion criteria | Published in English<br>Studied children with SAM | Studied patients had heart function assessed by ultrasound | Studied patients with cholera or diarrhea and severe dehydration +/- shock |
| Exclusion criteria | Utilized animal models or in-vitro data<br>Focused on MAM, and did not perform a sub analysis for children with SAM | -- | -- |

Data were abstracted by one author (RN) and cross checked by an independent author (KDT).

### Review of baseline cardiac function

Studies that evaluated cardiac function were assessed. Three areas of cardiac function were included: 1) Left ventricular mass, 2) Systolic function including ejection fraction and fractional shortening, and 3) Diastolic function, including grades of diastolic dysfunction and end diastolic diameter. Global cardiac function measures were also interpreted including the Tei or myocardial perfusion index (MPI).

### Review of intravenous rehydration

Studies that evaluated intravenous rehydration in children with SAM and cholera or diarrheal disease with severe dehydration with or without shock were assessed. Two main issues were addressed regarding current GTFCC guidance:

- Volume and infusion rates of intravenous rehydration given during resuscitation
- Utilization of isotonic versus hypotonic or hypertonic intravenous fluids during resuscitation

The following outcomes were considered in this section of the review: the incidence of fluid overload or heart failure, mortality and adverse events associated with different regimens, treatment failure, and evidence of depressed cardiac function utilizing cardiac ultrasound during intravenous rehydration. The study location, definition of SAM, years of operation, fluid regimens used, and proportion of kwashiorkor.

Data were abstracted from the included studies by a single author (RN) and verified by a second author (KDT) using standardized templates. Because of the limited number of studies identified and the heterogeneous populations and methods employed across the studies, meta-analysis was not attempted. The results are presented as a narrative review.

A modified GRADE approach was taken to assess the certainty of evidence. Risk of bias was assessed in all studies. For the first search, which included cross sectional assessments of baseline cardiac function the Joanna Briggs Institute (JBI) criteria for assessing cross sectional studies were used. For the second search, which assessed response to intravenous fluids, the Cochrane collaboration Risk of Bias 2 (ROB2) tool was adapted and employed. All risk of bias and certainty of evidence summaries were conducted by one author (KDT). The protocols for these reviews were not published prior to conducting the analysis.

## Results

Of 1813 records identified in the first search, eight studies assessing cardiac function in children with SAM were included (Figure 1a). The search assessing intravenous rehydration among children with SAM and severe dehydration with or without shock, included three out of 470 identified studies after screening and evaluation (Figure 1b).

**Figure 1:**
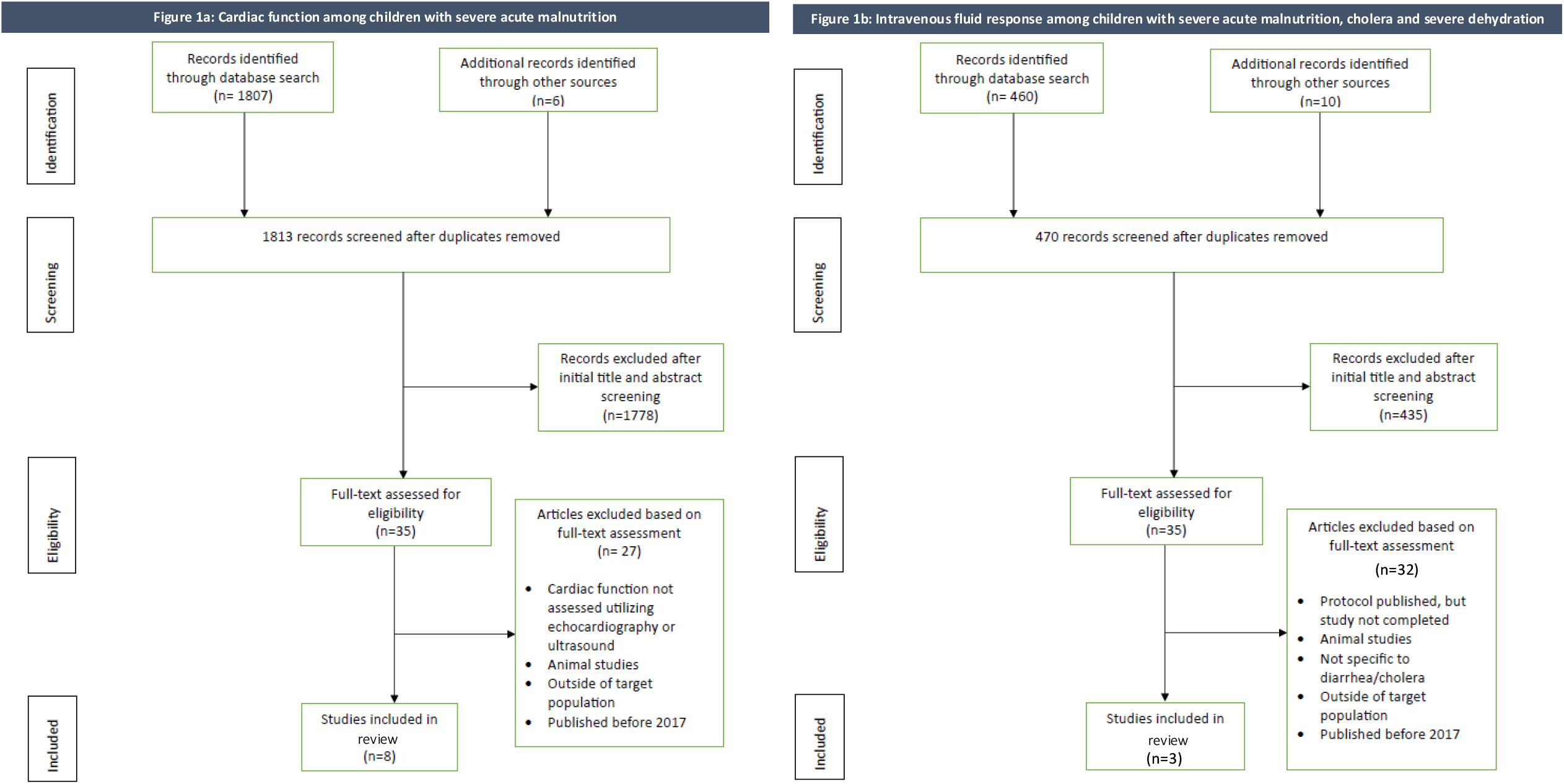
Study selection for the reviews of cardiac function and response to intravenous fluids.

### Baseline cardiac function among children with SAM

#### Description of Included Studies

Eight studies assessed cardiac function prior to fluid rehydration in children with SAM (S1 Appendix 2).[13,14,17–22] Two of the studies also assessed the response to intravenous rehydration review: The Appropriate Fluid Resuscitation in Malnutrition (AFRIM) study,[13] a prospective observational study, conducted in Kenya and Uganda and the Cardiac Physiology in Malnutrition (CAPMAL) study,[14] a prospective, matched case-control study, conducted in Kenya. In addition to AFRIM and CAPMAL, four studies were conducted in India, one was conducted in Pakistan, and one in Egypt. The eight studies were observational studies and used the WHO criteria to define SAM. AFRIM used hypovolaemic shock as an inclusion criteria.

Systolic function refers to the heart’s ability to contract and pump blood to the rest of the body, whereas diastolic function refers to the relaxation and filling phase of the heart. All studies used cardiac ultrasound to assess these parameters. All eight studies evaluated ejection fraction and fractional shortening, the primary parameters used to assess systolic function. Seven studies assessed left ventricular mass. Assessment of diastolic function is more complex in children and uses multiple different grading systems.[23] Direct assessment of diastolic function was performed in one study, but the other seven studies included indirect parameters that relate to diastolic function, including end diastolic diameter or the Tei/MPI which offers a global assessment of cardiac function.

#### Systolic Function and Left Ventricular Mass

914 children with SAM underwent cardiac ultrasound including 126 children with oedematous malnutrition. None (0/8) of the studies found decreased systolic function in SAM children compared to children without acute malnutrition or reference values. All seven studies reporting left ventricular mass found it to be significantly decreased in children with SAM. However, these seven studies also found preserved systolic function despite the lower left ventricular mass. No studies comparing the clinical phenotypes of severe wasting to oedematous malnutrition found decreased systolic function, although one study noted significantly lower left ventricular mass among children with nutritional oedema.

#### Diastolic Function

Results of seven of the included studies were consistent with decreased diastolic function among children with SAM. Bebars et al found that 30% of their 60 children with SAM had Grade II diastolic dysfunction, while another 40% had Grade I diastolic dysfunction, and the remaining 30% had normal function. None of the 120 children without SAM in that study were found to have diastolic dysfunction. The authors also found that after nutritional rehabilitation, 3% of children still had Grade II, 31% had Grade I, and 66% had no diastolic dysfunction.

Three studies measured end diastolic diameter and found it to be reduced among children with SAM compared to children without acute malnutrition. This is consistent with a transient diastolic dysfunction where the ventricular filling is restricted by impaired relaxation. Similarly, three of four studies included Tei/MPI global assessments of cardiac function and found it to be abnormally raised among children with SAM potentially indicating diastolic dysfunction. Opondo et al, defined a MPI >0.4 to be dysfunctional, and prior to rehydration found three (15%) of the 20 children with SAM were above this cut off. Jain et al found the mean MPI among 76 children with SAM to be 0.70 (standard deviations (SD): 0.10) compared to 0.43 (SD: 0.04) among 76 children without severe wasting. Similarly, Arshad et al also found a mean MPI of 0.72 (SD: 0.12) among children with SAM and 0.45 (SD: 0.04) among control children. However, Brent et al found 88 children with SAM to have a normal mean Tei/MPI index (0.37), but significantly different E/E’ ratios between children with oedematous and severe wasting. No studies noted differences in diastolic function or Tei/MPI between children with severe wasting only and those with oedematous malnutrition.

#### Certainty of Evidence

The certainty of evidence for changes in left ventricular mass, systolic function, and diastolic function are given in Table 2. The evidence is moderately certain that children with SAM have a reduced left ventricular mass but their systolic function remains comparable to better nourished children. We have a low certainty in the evidence regarding diastolic dysfunction, because there is inconsistency (one study with a low risk of bias found no diastolic dysfunction). Additionally, measures of end diastolic diameter could be influenced by dehydration and acute illness suggesting a risk of bias.

**Table 2:**
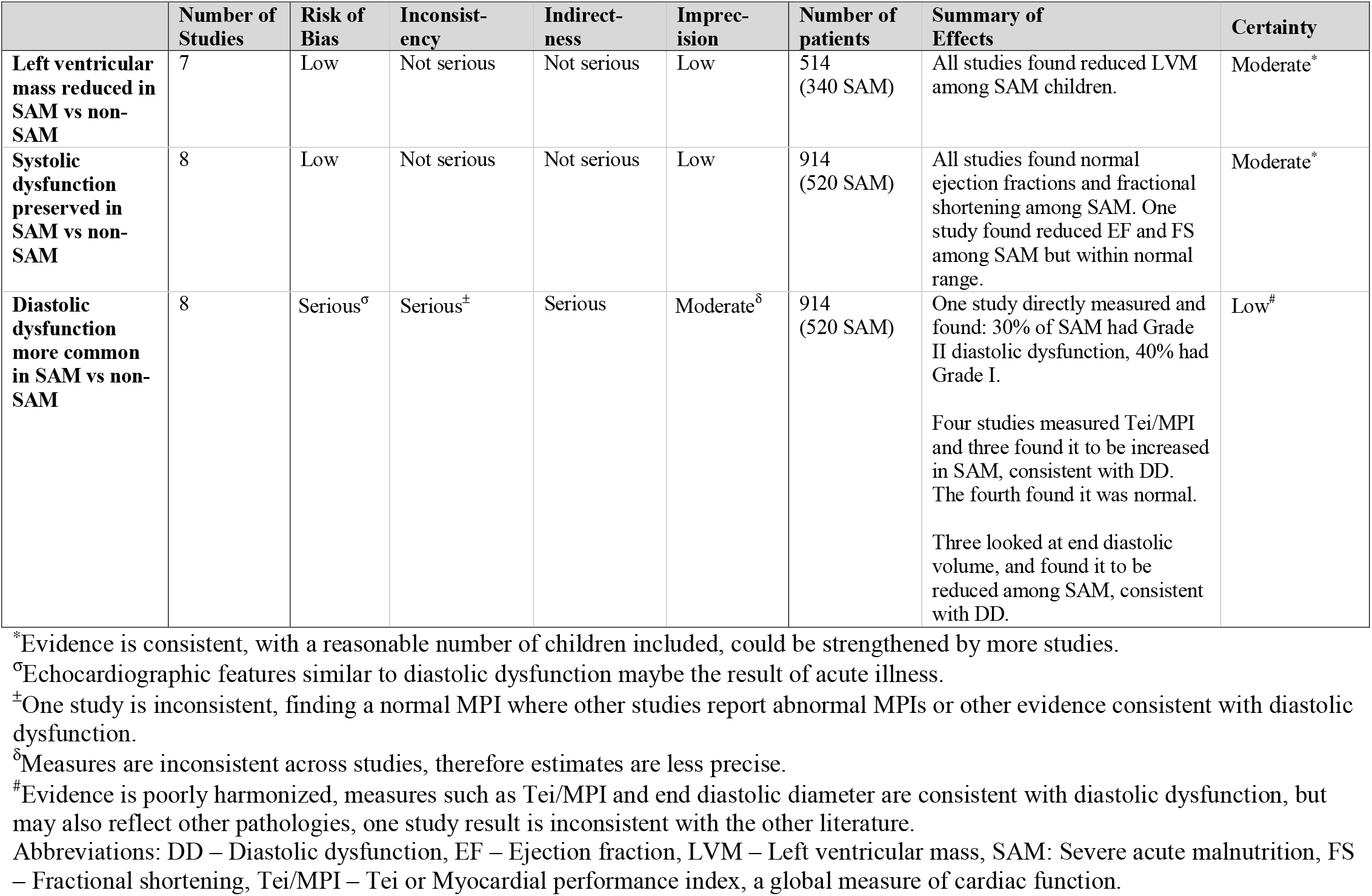
Summary of evidence for cardiac function among children with SAM compared to those without SAM.

|  | Number of Studies | Risk of Bias | Inconsistency | Indirectness | Imprecision | Number of patients | Summary of Effects | Certainty |
| --- | --- | --- | --- | --- | --- | --- | --- | --- |
| **Left ventricular mass reduced in SAM vs non-SAM** | 7 | Low | Not serious | Not serious | Low | 514 (340 SAM) | All studies found reduced LVM among SAM children. | Moderate* |
| **Systolic dysfunction preserved in SAM vs non-SAM** | 8 | Low | Not serious | Not serious | Low | 914 (520 SAM) | All studies found normal ejection fractions and fractional shortening among SAM. One study found reduced EF and FS among SAM but within normal range. | Moderate* |
| **Diastolic dysfunction more common in SAM vs non-SAM** | 8 | Serious<sup>σ</sup> | Serious<sup>±</sup> | Serious | Moderate<sup>δ</sup> | 914 (520 SAM) | One study directly measured and found: 30% of SAM had Grade II diastolic dysfunction, 40% had Grade I.<br>Four studies measured Tei/MPI and three found it to be increased in SAM, consistent with DD. The fourth found it was normal.<br>Three looked at end diastolic volume, and found it to be reduced among SAM, consistent with DD. | Low<sup>#</sup> |

\*Evidence is consistent, with a reasonable number of children included, could be strengthened by more studies.
<sup>σ</sup>Echocardiographic features similar to diastolic dysfunction maybe the result of acute illness.
<sup>±</sup>One study is inconsistent, finding a normal MPI where other studies report abnormal MPIs or other evidence consistent with diastolic dysfunction.
<sup>δ</sup>Measures are inconsistent across studies, therefore estimates are less precise.
<sup>#</sup>Evidence is poorly harmonized, measures such as Tei/MPI and end diastolic diameter are consistent with diastolic dysfunction, but may also reflect other pathologies, one study result is inconsistent with the other literature.
Abbreviations: DD – Diastolic dysfunction, EF – Ejection fraction, LVM – Left ventricular mass, SAM: Severe acute malnutrition, FS – Fractional shortening, Tei/MPI – Tei or Myocardial performance index, a global measure of cardiac function.

### Response to intravenous rehydration among children with SAM and cholera or diarrheal disease plus severe dehydration with/without shock

#### Description of Included Studies

Three studies assessed children with SAM and diarrheal disease plus severe dehydration with or without shock who received intravenous rehydration (S1 Appendix 3).[13,14,16] All three studies utilized the WHO SAM case definition, and all their interventions used isotonic fluid preparations during initial management.

In the AFRIM study described above, the first 11 children with SAM and shock were given the WHO recommended management for shock beginning with up to two boluses of 15mL/kg/hour of intravenous Ringer’s Lactate and progressing to 4 mL/kg/hour of half-strength Darrow’s/5% dextrose. The next nine children were given a “rehydration-only” approach, including 10 mL/kg/hour for a maximum of 5 hours of intravenous Ringer’s Lactate without fluid boluses. Children in both groups were switched to rehydration solution for malnutrition, (ReSoMal) once oral or nasogastric fluids were tolerated. The children’s response to these regimens were monitored using echocardiography and regular assessment for clinical signs of fluid overload. Of the 20 children included in AFRIM, 15 had oedematous malnutrition, but the authors did not comment on differences in the below outcome by nutritional oedema status.

During the CAPMAL study, 12 children with SAM, gastroenteritis, and evidence of shock were given boluses of intravenous Ringer’s Lactate at 20 mL/kg/hour, repeated up to two times if ≥2 signs of shock remained without signs of pulmonary oedema. Echocardiography was conducted at study enrolment and during intravenous rehydration. Thirty-six of the 88 children with SAM in this study had oedematous malnutrition, but oedematous malnutrition was not discussed in relation to the below outcomes. Eleven of 14 deaths occurred after day 7.

Alam et al. conducted a randomized controlled trial in Bangladesh of rapid compared to slow rehydration in children with SAM and dehydrating diarrhea. V. cholerae was identified in 124/208 (60%) of the children [24]. The rapid hydration arm (n=105) included 100mL/kg of intravenous cholera saline (an isotonic fluid) given over 6 hours, with 30 mL/kg in the first 30 minutes, then 70 mL/kg over 5.5 hours. The slow rehydration arm was the WHO SAM protocol for shock of 15 mL/kg of intravenous fluid over 1 hour, and if respiratory and pulse rate improved another 15 mL/kg in the 2 hour followed by rice-based oral rehydration solution (ORS) based on estimated deficit. In addition to the intravenous regimen, both groups received ORS for ongoing stool loss. Nutritional oedema was present in 22-25% of this trial’s included children, but the authors did not comment on difference in outcome by nutritional oedema status.

#### Fluid Overload

The three included studies documented no cases of fluid overload among the 240 children with SAM enrolled. This included 117 children who were given intravenous fluids above the WHO recommended volumes in the Alam (2020) and CAPMAL studies.

#### Mortality

The AFRIM study reported 14/20 (70%) children died within 28 days, including nine of 11 (82%) children who received the WHO rehydration protocol, and five of nine children on “rehydration-only” protocol (56%). Four deaths occurred in each group in the first 24 hours. The difference between the two groups was not statistically significant, although the study was not powered to detect differences in mortality. Deaths were attributed to hypovolemic cardiovascular collapse, sepsis, respiratory arrest, aspiration, metabolic derangements/acidosis, multiorgan failure, or underlying comorbidities. No deaths were attributed to fluid overload. Alam et al. reported no deaths among the 208 children randomized to rapid or slow rehydration. CAPMAL is not included in our mortality summary as it did not compare regimens.

#### Treatment Success/Failure

In Alam 2020, a similar number of children failed to complete both the rapid and slow intravenous treatment regimens. No treatment failures were reported to be related to complication of the assigned treatment approach. Reported reasons for treatment failure included: convulsions, severe pneumonia, sepsis, hypokalaemia, and prolonged diarrhea. There was no difference in the output of vomit and stool between the groups, but the rapid rehydration group had significantly increased urine output during the initial 12 hours when compared to the slow rehydration group. The difference in volume of urine during the first 12 hours was not reported.

The AFRIM study noted that four of 11 (36%) children in their fluid bolus group and three of nine (33%) in their slow rehydration group required blood transfusions due to ongoing shock. Urine outputs of >1mL/kg/hour, deemed by the authors to be satisfactory, were noted in eight of 11 (73%) children in the fluid bolus group, and seven of nine (78%) in the slow rehydration group.

#### Response to Fluids Assessed by Echocardiography

The AFRIM and CAPMAL studies encompassed 32 children with SAM and signs of shock. Both studies found a heterogenous response to fluids, but concluded that the majority of children with SAM had an appropriate response to intravenous fluids indicated by increased stroke volumes and decreased heart rates. The AFRIM study observed at least one of their 20 children to be fluid non-responsive, where the child’s stroke volume index decreased, and their end diastolic volume increased after intravenous fluids. Similarly, CAPMAL observed three episodes where the stroke volume index decreased after intravenous fluids, but the investigators assessed that these were attributable to ongoing septic shock. Despite the heterogeneity of responses to intravenous fluids, none of the children developed clinical evidence of fluid overload.

#### Certainty of Evidence

The certainty of evidence regarding the incidence of fluid overload among children with SAM was judged to be low (Table 3). The evidence base is weakened by the non-randomized nature of two of the three studies, which leaves them at a high risk of bias. The Alam (2020) randomized control trial had a low risk of bias, but it cannot exclude fluid overload from being a rare adverse event. For both mortality and the echocardiographic assessment of response to fluids, the certainty of evidence was very low, because the sample sizes were small relative to the possible incidence of the outcome, and the results are highly heterogenous. The findings related to treatment success or failure have a very low certainty of evidence because the estimates lack precision.

**Table 3:**
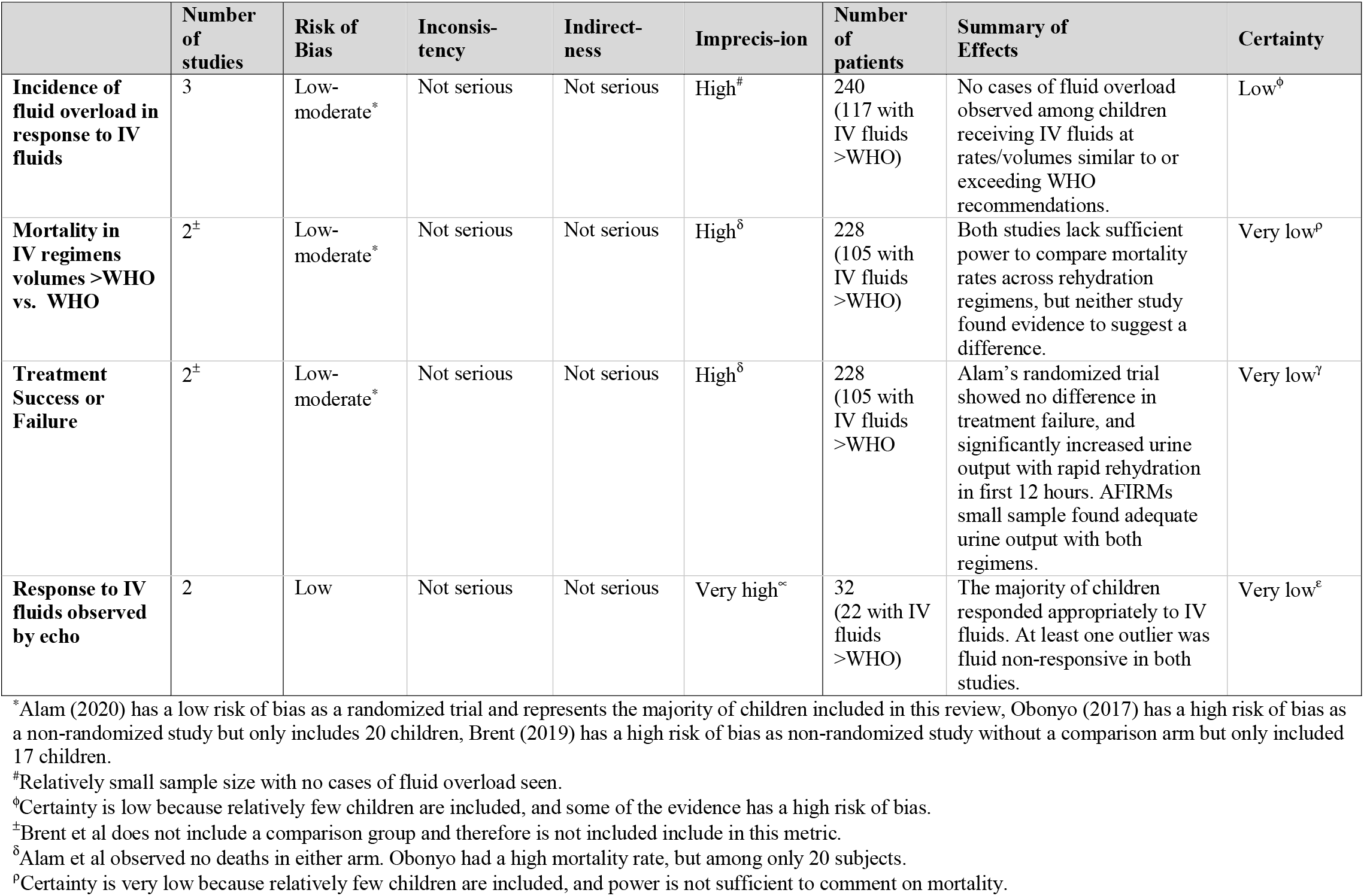
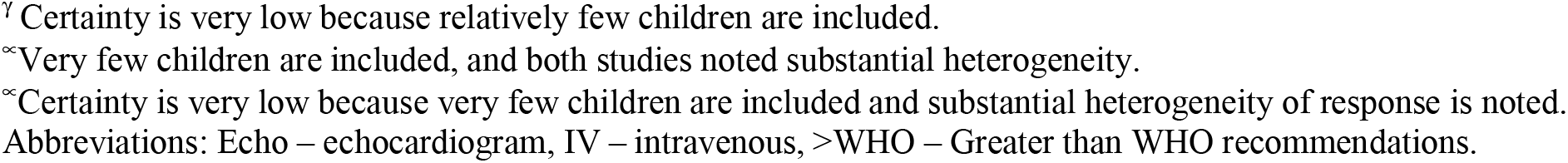
Summary of evidence for the safety of intravenous rehydration among children with SAM.

|  | Number of studies | Risk of Bias | Inconsistency | Indirectness | Imprecision | Number of patients | Summary of Effects | Certainty |
| --- | --- | --- | --- | --- | --- | --- | --- | --- |
| **Incidence of fluid overload in response to IV fluids** | 3 | Low-moderate<sup>*</sup> | Not serious | Not serious | High<sup>#</sup> | 240 (117 with IV fluids >WHO) | No cases of fluid overload observed among children receiving IV fluids at rates/volumes similar to or exceeding WHO recommendations. | Low<sup>φ</sup> |
| **Mortality in IV regimens volumes >WHO vs. WHO** | 2<sup>±</sup> | Low-moderate<sup>*</sup> | Not serious | Not serious | High<sup>δ</sup> | 228 (105 with IV fluids >WHO) | Both studies lack sufficient power to compare mortality rates across rehydration regimens, but neither study found evidence to suggest a difference. | Very low<sup>ρ</sup> |
| **Treatment Success or Failure** | 2<sup>±</sup> | Low-moderate<sup>*</sup> | Not serious | Not serious | High<sup>δ</sup> | 228 (105 with IV fluids >WHO) | Alam's randomized trial showed no difference in treatment failure, and significantly increased urine output with rapid rehydration in first 12 hours. AFIRMs small sample found adequate urine output with both regimens. | Very low<sup>γ</sup> |
| **Response to IV fluids observed by echo** | 2 | Low | Not serious | Not serious | Very high<sup>∞</sup> | 32 (22 with IV fluids >WHO) | The majority of children responded appropriately to IV fluids. At least one outlier was fluid non-responsive in both studies. | Very low<sup>ε</sup> |

<sup>\*</sup>Alam (2020) has a low risk of bias as a randomized trial and represents the majority of children included in this review, Obonyo (2017) has a high risk of bias as a non-randomized study but only includes 20 children, Brent (2019) has a high risk of bias as non-randomized study without a comparison arm but only included 17 children.
<sup>#</sup>Relatively small sample size with no cases of fluid overload seen.
<sup>φ</sup>Certainty is low because relatively few children are included, and some of the evidence has a high risk of bias.
<sup>±</sup>Brent et al does not include a comparison group and therefore is not included in this metric.
<sup>δ</sup>Alam et al observed no deaths in either arm. Obonyo had a high mortality rate, but among only 20 subjects.
<sup>ρ</sup>Certainty is very low because relatively few children are included, and power is not sufficient to comment on mortality.
<sup>γ</sup> Certainty is very low because relatively few children are included. <sup>∞</sup>Very few children are included, and both studies noted substantial heterogeneity. <sup>∞</sup>Certainty is very low because very few children are included and substantial heterogeneity of response is noted. Abbreviations: Echo – echocardiogram, IV – intravenous, >WHO – Greater than WHO recommendations.

## Discussion

The studies including assessments of cardiac function suggest that children with SAM do not show evidence of cardiac systolic dysfunction despite decreased left ventricular mass. This observation was consistent across the included studies, and echoes the conclusions of a systematic review published in 2017.[25] We found some evidence of a transient diastolic dysfunction among a subset of children with SAM, but the clinical significance of this reduced diastolic function is unclear. We did not find any documented cases of fluid overload during intravenous rehydration of children with SAM and severe dehydration.

Diastolic dysfunction may explain the discrepancy between the long-held belief that children with SAM are vulnerable to fluid overload despite consistent evidence of preserved systolic function relative to their body mass. Children with Grade II diastolic dysfunction may have raised left atrial pressures, and therefore a theoretically lower threshold for fluid overload.[26] However, diastolic dysfunction is not a contraindication to fluid resuscitation among children. A recent survey of 387 pediatric intensivists from 48 countries found some degree of equipoise between the 58% that would fluid restrict while resuscitating children with proven cardiac dysfunction and the 42% who would not.[27] The AFRIM and CAPMAL studies found some evidence that children with SAM may have a heterogeneous response to fluids, but noted that none of the children developed fluid overload.[13,14] Among the 20 children in AFRIM, at least six had increased brain natriuretic peptide or echocardiographic evidence of heart strain at baseline, but only one child responded poorly to intravenous fluids. This suggests that among the subset of children with SAM who have some evidence of cardiac dysfunction there could be a further subset who may respond poorly to intravenous fluids.

Combining our findings with the 2017 review,[25] at least 476 children with SAM have received intravenous rehydration as part of a published clinical study and none have been found to have evidence of fluid overload. These data are further strengthened by the recently published GastroSAM trial in which 134 young children with SAM were given intravenous fluids (67 rapid infusion, 67 slow infusion) without any evidence of fluid overload.[28] We cannot exclude fluid overload as a rare event, and the evidence lacks sufficient power to meaningfully comment on mortality differences between regimens. However, it does suggest fluid overload is not a common outcome for children with SAM and severe dehydration when given intravenous fluids. We also note that the WHO pocket book provides guidance on the management of fluid overload without requiring access to higher levels of pediatric intensive care or ventilation.[4] The existing evidence does not exclude a theoretical risk of fluid overload, a severe but manageable adverse event, in a subset of patients. The risk of fluid overload among a subset of children with SAM must be weighed against the very high purging rates cholera patients often experience. Severe cholera can result in approximately 100mL/kg of fluid loss in 24 hours, far exceeding purging rates of many other diarrheal diseases, which can rapidly lead to hypovolemia, severe dehydration and shock in children.[3] These findings suggest that we should re-examine policies limiting intravenous resuscitation in children with SAM and severe dehydration from cholera.

Alam 2020 showed some evidence that rapid intravenous rehydration of children with SAM increased urine output in the first 12-hours of treatment.[16] The WHO pocket book notes that urine output is the most sensitive marker of fluid status among children. This may suggest that the rapid intravenous rehydration utilized by Alam et al restored urine output and corrected dehydration faster than a more cautious approach, and supports the argument that intravenous rehydration may benefit children with SAM and cholera.

One of the major limitations in this literature is that much of the critical evidence is drawn from children with shock, which may not generalize to children with severe dehydration but without shock. This is particularly evident in the causes of death in the AFRIM study, and the heterogeneity of intravenous fluid responsiveness in AFRIM and CAPMAL. In both these studies, the investigators concluded that several mortalities, and the cases of fluid non-responsiveness, were caused by on-going sepsis and not fluid overload.[13,14] These findings echo the conclusions of the FEAST trial, in which children without acute malnutrition who were in shock and given fluid boluses were found to be at increased risk of mortality, but these deaths were attributed to cardiovascular collapse and not fluid overload.[29] Alam et al (2020), excluded children with sepsis and did not see the high mortality rates observed in AFRIM and CAPMAL.

Isotonic fluids are recommended for intravenous rehydration of hypovolemic children over hypotonic or hypertonic solutions, for rapid volume expansion and a lower risk of electrolyte shifts. However, there should be close monitoring and correction of hypoglycemia for children with SAM.[4] All the studies included in this review used an isotonic fluid, but did not comment on the management of hypoglycaemia. The included studies do not offer conclusive evidence about the optimal rate of fluids for children with SAM and severe dehydration, but the recent GastroSAM trial gave 100mls/kg over the first three-to-six hours, similarly Alam (2009) gave 100ml/kg in six hours, while Alam (2020) gave 60mls/kg in three hours. Frequency of monitoring is also likely to be a critical determinant of safe fluid rehydration. WHO recommends children with SAM and shock are monitored every five minutes during rehydration, but the feasibility of implementing these frequencies in busy centers must be considered. Interestingly Alam (2020) reports that they only monitored once per hour.

This review is limited by the current evidence base which lacks large, definitive randomized trial evidence. The randomized controlled evidence available is largely from a single institution in Bangladesh, although the publication of GastroSAM broadens this evidence base. Other available evidence has focused on the management of shock, which should be evaluated separately for severe dehydration without shock. Analyses of mortality are severely under powered, and the echocardiography studies have small sample sizes. Finally, some observations, such as reduced end diastolic volume and increased MPI may be caused by other features of SAM and diarrhea, for example reduced muscle mass and dehydration.

This review reaffirms previous findings that children with SAM have comparable cardiac systolic function to children without SAM despite having lower left ventricular mass, and that studies of intravenous rehydration among children with SAM have not reported cases of fluid overload. We did note some evidence of lower-grade, transient, diastolic dysfunction among a subset of children with SAM. The potential for dysfunction among a subset of children with SAM reinforces the need for close monitoring during intravenous rehydration and institutional capacity to manage fluid overload. However, the majority of children with SAM appear to be cardiologically competent and may benefit from intravenous fluid resuscitation for severe dehydration, especially in conditions with high purging, such as cholera.

## Data Availability

All data produced in the present work are contained in the manuscript

## Acknowledgements

This review was conducted with support from the GTFCC Case Management Working Group. We would like to also like to acknowledge the financial support provided from the United States Government Bureau of Humanitarian Affairs (USAID/BHA). A special thanks to Dr Alam from the International Centre for Diarrhoeal Disease Research, Bangladesh for his time and expertise dedicated to this review.

