## Appendices for "Systematic review of the evidence of cardiac dysfunction and intravenous fluid use among children with severe acute malnutrition and severe dehydration"

**S1 Appendices**

**Appendix 1**: Comparison of IV rehydration protocols for children with SAM and cholera or diarrheal disease with severe dehydration.

| Condition | **World Health Organization^[1]^** | **The Global Task on Cholera Control^[2]^** | **Médecins Sans Frontières (MSF) ^[3]^** | **International Centre for Diarrheal Disease Research, Bangladesh^[4]^** |
| --- | --- | --- | --- | --- |
| SAM with severe dehydration without shock | Standard ORS, 5mL/kg every 30 mins for 2 hours. Then 5-10mLs/kg/hr alternating with F75. give an additional 50mLs after loose stool if < 2YO. 100mLs if > 2YO | Standard ORS, 5mL/kg every 30 mins for 2 hours. Then 5-10mLs/kg/hr alternating with F75. Give an additional 50 mLs after loose stool if < 2YO. 100mLs if > 2YO | 20 mL/kg IV bolus over 30 minutes with RL.  Repeat same 20 mL/kg bolus (up to 2 times) if danger signs still present  **If danger signs resolve: 70 mL/kg of RL over 6 hours; continue glucose adding** 100mL of 50% glucose to each liter of RL) | 20 mL/kg IV bolus + match purging over 1^st^ hour  20 mL/kg + match purging (10 mL/kg IV + 10mL/kg ORS) over 2^nd^ hour  Use Full strength acetate (cholera saline) with 5% dextrose with or without 7 mmol/L KCL (for >2 months of age) |
| SAM with Shock | 15 mL/kg IV bolus in first hour with  HS Darrow’s plus 5% dextrose, OR RL with 5% dextrose. 0.45% saline with 5% dextrose if neither available.  Repeat bolus in second hour if improved. If shock not improved after 1 hour of IV fluid, give whole blood at 10 mL/kg slowly over at least 3 hours | 15 mL/kg IV bolus over 1 hour with  Half-strength Darrow’s solution plus 5% dextrose, OR RL with 5% dextrose. 0.45% saline with 5% dextrose if neither available.  If child does not improve after 1 hour of rehydration, assume septic shock and treat accordingly | 20 mL/kg IV bolus over 30 minutes with RL.  Repeat same 20 mL/kg bolus (up to 2 times) if danger signs still present  **If danger signs resolve: 70 mL/kg of RL over 6 hours; (continue glucose by adding** 100 mL of 50% glucose to each liter of RL) | 20 mL/kg IV bolus + match purging over 1^st^ hour  20 mL/kg + match purging (10 mL/kg IV + 10mL/kg ORS) over 2^nd^ hour  Use Full strength acetate (cholera saline) with 5% dextrose with or without 7 mmol/L KCL (for >2 months of age)  After 2 hours, IV is discontinued and ORS is given at 10 mL/kg/hour for next 2 hours |

**Appendix 2:** Summary of the literature evaluating cardiac function in children with SAM

| **Author, Year** | **Country** | **Study type** | **Population** | **Inclusion criteria** | **Exclusion criteria** | **Decreased Left Ventricular Mass**  **on echocardiography** | **Normal systolic function on echocardiography** |
| --- | --- | --- | --- | --- | --- | --- | --- |
| Arshad et al., 2022 | Pakistan | Prospective | N=150  75 SAM  26/75, 35% Kwashiorkor  75 non-SAM controls | WHO SAM criteria | Not specified | 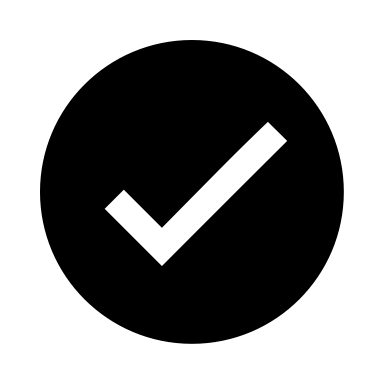 | 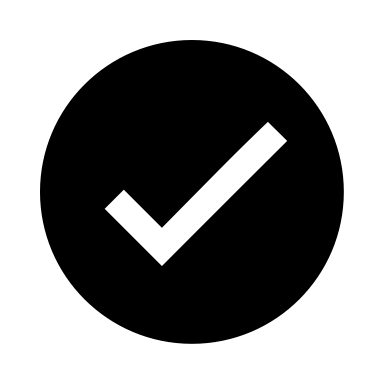 |
| Bebars & Askalany, 2019 | Egypt | Follow-up case control | N=180  60 SAM  120 non-SAM controls | WHO SAM criteria | -history of cardiothoracic event  -congenital heart disease  -metabolic or glycogen storage disease  -chronic medical disease  -chronic liver disease  -prematurity or interuterine growth restrcition.  -muscle disease  -diabetes | *Study did not assess this parameter* | 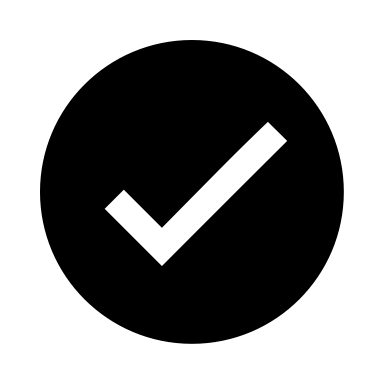 |
| Brent et al., 2019 | Kenya | Prospective, matched, case-control | N=110  88 SAM  36/88, 41% Kwashiorkor  22 non-SAM controls | WHO SAM criteria | -congenital heart disease | 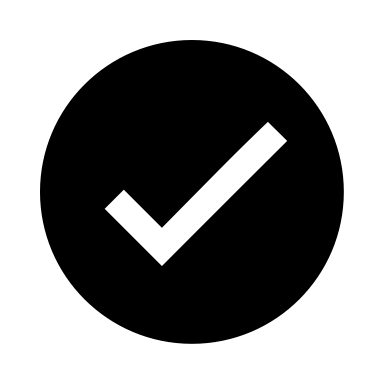 | 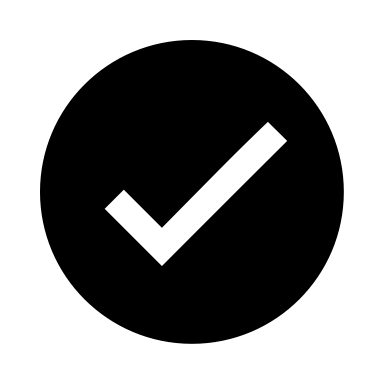 |
| Jain et al., 2019 | India | Prospecitve case-control | N=152  76 SAM  49/76, 64% Kwashiorkor  76 non-SAM controls | WHO SAM criteria | -prematurity or IUGR  -inborn error of metabolism  -congential anomalies  -chronic renal failure  -cerebral palsy  -chronic liver disease  -chromosomal abnormalities | 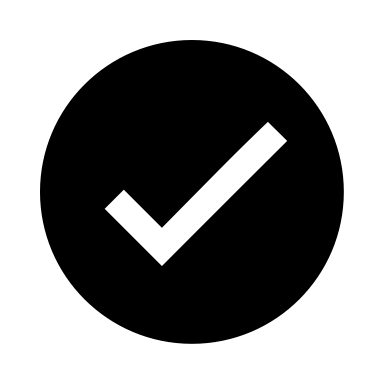 | 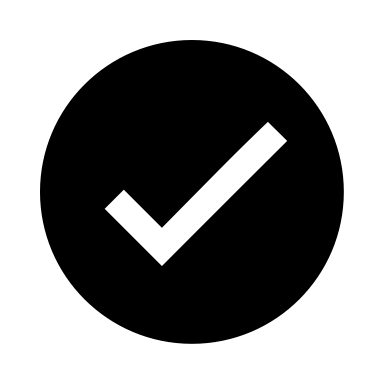 |
| Obonyo et al., 2017 | Kenya  Uganda | Prospective observational | N =20 SAM  15/20, 75% Kwashiorkor | WHO SAM criteria  **PLUS** acute hypovolemic diarrhea  **WITH** signs of severe dehydration and shock | -severe dermatitis of groin  - congenital heart disease | 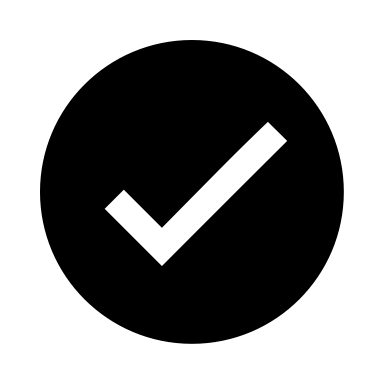 | 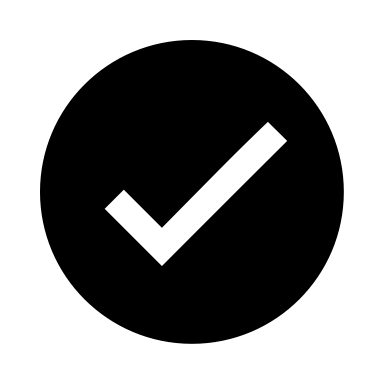 |
| Pradhaa HMV et al, 2023 | India | Cross sectional observational | N=50 SAM | WHO SAM criteria | -congenital or acquired heart disease  -prematurity  -IUGR  -congenital anomalies  -systemic or chronic disease | 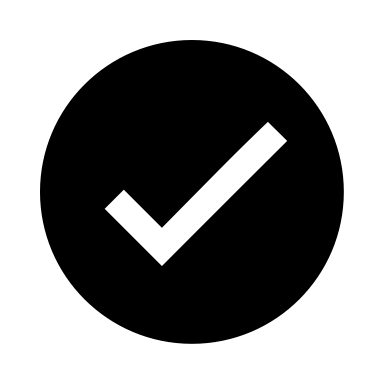 | 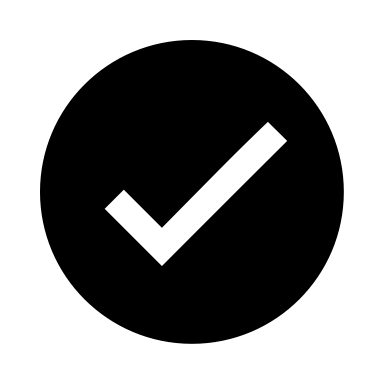 |
| Samikannu et al, 2019 | India | Case-control | N=62  41 SAM  21 non-SAM controls | WHO SAM criteria | -prematurity or interuterine growth restriction  -congenital heart disease  -acquired heart disease  -severe anemia | 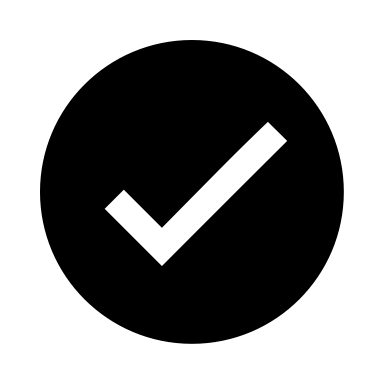 | 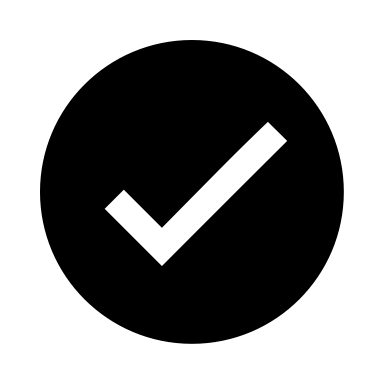 |
| Sharma et al., 2017 | India | Prospective case-control | N=200  100 SAM  100 non-SAM controls | WHO SAM criteria | Not specified | 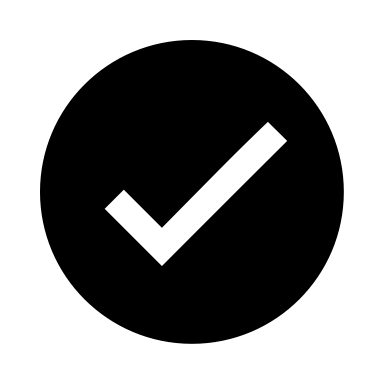 | 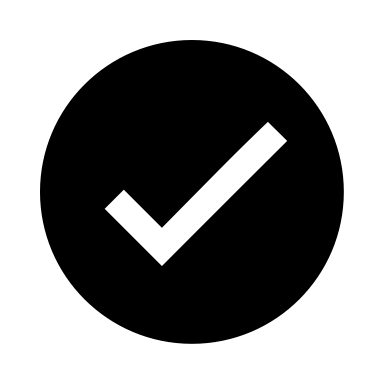 |

**Appendix 3:** Table showing methods and results/conclusions from studies investigating rehydration in children with SAM, diarrheal disease and severe dehydration +/- shock

| **Author, Year** | **Location/**  **Study type** | **Study Type** | **Population** | **Sample size** | **Inclusion** | **Exclusion** | **Outcomes** |
| --- | --- | --- | --- | --- | --- | --- | --- |
| Alam et al., 2020 | Bangladesh  ICDDR,B | RCT | Children 6-36 months with SAM, acute diarrheal disease and severe dehydration | 208 | SAM defined as:  weight for age and weight for length Z-scores <-3 with or without bipedal edema  Acute diarrhea defined as ≥3 loose or watery stools in <24 hours  Severe dehydration defined by modified WHO guidelines | - Dysentery - Severe pneumonia - Suspected severe sepsis or septic shock - Suspected meningitis - Antibiotic use before admission | - Required unscheduled IV therapy - Duration of diarrhea after admission - Electrolyte abnormalities after 24 hours - Fluid overload |
| Obonyo et al., 2017 | Kenya, Uganda | Cohort | Children 6-60 months with SAM, with diarrhea and severe dehydration and shock | 20 | SAM defined using WHO criteria  Hypovolemic diarrhea: >3 watery stools/24 hours  Severe dehydration and shock (2 or more signs below):  -Cap refill ≥3 seconds  - Temp gradient between extremities and core body  - rapid and/or weak pulse volume | - Severe dermatitis of groin - Congenital heart disease | - Data collected on assessment of myocardial function and hemodynamic response with fluid resuscitation using echocardiography |
| Brent et al, 2019 | Kenya | Cohort | Children with SAM and shock | 12  (with shock) | SAM defined using WHO criteria and shock:  -Cap refill ≥3 seconds  - Temp gradient between extremities and core body  - Rapid and/or weak pulse volume  -Lethargic or unconscious | - Congenital heart disease | - Echogram assessment of response to intravenous fluids |

**Annex 5** Comparison of IV fluid composition^[5]^

| **IV Solution** | **Na+** | **K+** | **Cl−** | **Acetate** | **Glucose** | **Tonicity** |
| --- | --- | --- | --- | --- | --- | --- |
|  | **mmol/L** | **mmol/L** | **mmol/L** | **mmol/L** | **g/L** |  |
| Ringer's lactate | 130 | 5.4 | 112 | 27 | – | Isotonic |
| Normal saline (0.9% NaCl) | 154 | – | 154 | – | – | Isotonic |
| 0.45% NaCl/5% glucose | 77 |  | 77 |  | 50 | Hypertonic |
| Ringer’s lactate with 5% glucose | 130 | 4 | 109 | 28 | 50 | Hypertonic |
| Half-strength Darrow with 5% glucose | 61 | 17 | 52 | 27 | 50 | Hypotonic |
| Cholera saline | 133 | 13 | 98 | 48 |  | Isotonic |
